# The effect of eye protection on SARS-CoV-2 transmission: a systematic review

**DOI:** 10.1101/2021.08.08.21261770

**Authors:** Oyungerel Byambasuren, Elaine Beller, Justin Clark, Peter Collignon, Paul Glasziou

## Abstract

**Background:** The effect of eye protection to prevent SARS-CoV-2 infection in the real-world remains uncertain. We aimed to synthesize all available research on the potential impact of eye protection on transmission of SARS-CoV-2.

**Methods:** We searched PROSPERO, PubMed, Embase, The Cochrane Library for clinical trials and comparative observational studies in CENTRAL, and Europe PMC for pre-prints. We included studies that reported sufficient data to estimate the effect of any form of eye protection including face shields and variants, goggles, and glasses, on subsequent confirmed infection with SARS-CoV-2.

**Findings:** We screened 898 articles and included 6 reports of 5 observational studies from 4 countries (USA, India, Columbia, and United Kingdom) that tested face shields, googles and wraparound eyewear on 7567 healthcare workers. The three before-and-after and one retrospective cohort studies showed statistically significant and substantial reductions in SARS-CoV-2 infections favouring eye protection with odds ratios ranging from 0.04 to 0.6, corresponding to relative risk reductions of 96% to 40%. These reductions were not explained by changes in the community rates. However, the one case-control study reported odds ratio favouring no eye protection (OR 1.7, 95% CI 0.99, 3.0). The high heterogeneity between studies precluded any meaningful meta-analysis. None of the studies adjusted for potential confounders such as other protective behaviours, thus increasing the risk of bias, and decreasing the certainty of evidence to very low.

**Interpretation:** Current studies suggest that eye protection may play a role in prevention of SARS-CoV-2 infection in healthcare workers. However, robust comparative trials are needed to clearly determine effectiveness of eye protections and wearability issues in both healthcare and general populations.

**Funding:** There was no funding source for this study. All authors had full access to all data and agreed to final manuscript to be submitted for publication.

## Introduction

Facial protection – for both wearer and close contacts – has been a crucial and controversial feature of the COVID-19 pandemic. For example, WHO recommends eye protection (goggles or face shield) for health care workers caring for COVID-19 patients but are not currently recommended for those caring for COVID-19 patients at home even when in the same room. A major uncertainty has been how much protection is provided by different forms and combinations of facial coverings.

We know that respiratory viruses such as SARS-CoV-2 can enter the respiratory tract via the nose, mouth, or eyes and inoculation may occur via air-to-face or hands-to-face. Early in the COVID-19 pandemic the mix of these routes was unclear, but now the air-to-face route is agreed to be an important factor in most cases. Less clear is what are the proportions of inoculation that occurs via the nose versus eyes. The cornea has ACE-2 receptors which may allow SARS-CoV-2 infection, but more likely is inoculation of the nasal epithelium via the nasolacrimal duct^1^.

During the Spanish influenza epidemic, evidence was presented in 1919 on the potential importance of eyes in the transmission of pathogens, and therefore using some form of eye protection^2^. Despite evidence from the previous SARS and MERS outbreaks suggesting an impact of eye protection^3^, the COVID-19 pandemic has seen much less research focused on eye protection. A call for face shields to provide eye protection early in the pandemic seemed to be largely ignored in both practice and research^4^, despite some promising studies. One early observational study in India of healthcare workers dealing with COVID-19 patients in the community showed a dramatic decline in the numbers of workers getting infected after face shields were made mandatory^5^. However, this study has received relatively little attention. A recent plea in the Lancet Microbe pointed to eye protection as a potential missing key^1^.

Therefore, to examine the potential contribution of eye protection, we aimed to identify, appraise, and synthesise all studies that estimated the impact of any form of eye protection including face shields and variants, goggles, glasses, and others on transmission of SARS-CoV-2.

## Method

We conducted a systematic review using enhanced processes and automation tools^6^. We searched the PROSPERO database to rule out existence of a similar review; searched PubMed, Embase, and The Cochrane Library’s CENTRAL for clinical studies, and Europe PMC for pre-prints from 1 Jan 2020 to 1 Jun 2021. A search string composed of MeSH terms and words was developed in PubMed and was translated to be run in other databases using the Polyglot Search Translator^7^. The search strategies for all databases are presented in Supplement 1. We also searched World Health Organization – International Clinical Trials Registry Platform (ICTRP) and ClinicalTrials.gov databases from inception until 1st January 2020. Protocol for this review was developed but not registered. We used PRISMA 2020 statement as a reporting guideline for this review.^8^

All publication types and languages were included in the search. We also conducted forward and backward citation searches for included studies in the Scopus citation database.

Our inclusion criteria – based on participants, interventions, and outcomes - were:

- Participants: all studies in humans, whether community or health care workers.
- Interventions: any form of eye protection, including face shields, goggles, or modified snorkel masks, with or without face masks.
- Comparators: No eye protection, with or without face masks.
- Outcomes: number of laboratory-confirmed infection with SARS-CoV-2.

We included any comparative study design (including before-and-after). We excluded studies if they did not provide sufficient data to make a comparison between eye protection and no eye protection; laboratory experiments; and any other eyewear that was not designed for prevention of respiratory virus transmission.

### Screening

Two authors (OB, EB) independently screened titles, abstracts, and full texts. Full text articles were retrieved by OB. Discrepancies were resolved by referring to a third author (PG). The selection process was recorded in sufficient detail to complete a PRISMA flow diagram (see Figure 1) and a list of excluded (full text) studies with reasons for exclusions are provided in Supplement 2.

**Figure 1.**
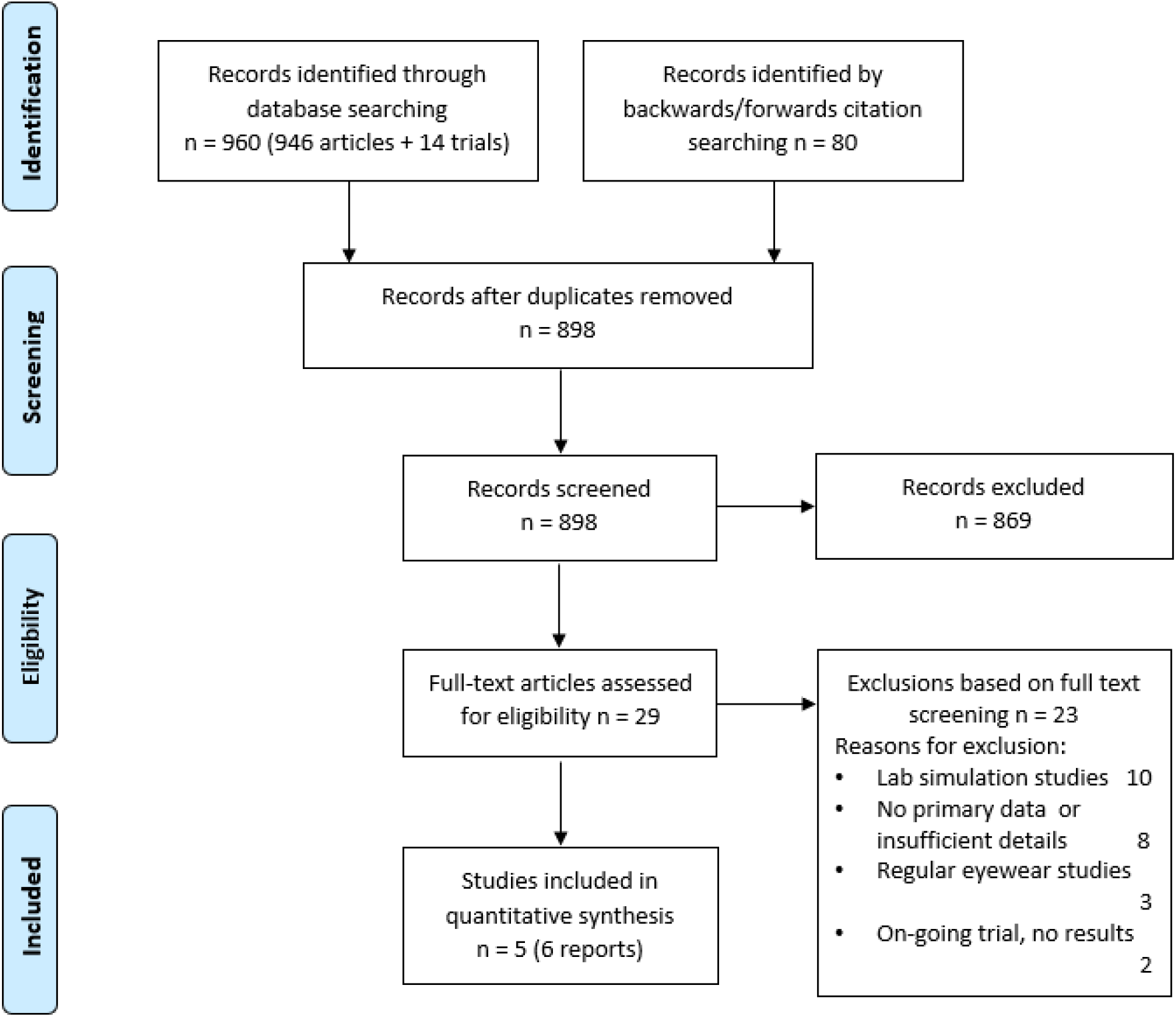
Screening and selection of articles.

### Data extraction

Two authors independently extracted the following data from included studies:

a. Study characteristics: country, study type, study setting, details of study PICO, timeframe,
b. Quantitative outcomes: total number of populations tested, total number of COVID(+) cases both before and after the intervention was mandated.
c. Number of cumulative COVID-19 cases in the community before and after the intervention, from external sources when necessary.

### Assessment of the Risk of bias

Two authors (OB, EB) independently assessed the risk of bias for each study using the ROBINS-I for observational studies^9^. We identified the following confounding domains relevant to most studies in order of most to least likely:

a. change in risk of COVID-19 from before, during and after the intervention,
b. test frequency or who is being tested (i.e., because of change in those being followed for infection),
c. comparator (use or compliance with other PPE),
d. setting (community, hospital, COVID-specific ward). Summary of the overall risk of bias for the 5 included studies is shown in Table 2.

We also identified the following two co-interventions that could be different between intervention groups and that could impact outcomes: (i) Addition of other infection control measures. Particularly important in before-after studies; (ii) Frequency of testing for COVID-19 (e.g., change in policy for testing, mandating of testing).

### Data analysis

We planned to do meta-analyses when two or more studies reported the same outcome provided that the heterogeneity was sufficiently low. The forest plot of intervention effects was created using Review Manager (RevMan) Version 5.4.1 (Copenhagen: The Nordic Cochrane Centre, The Cochrane Collaboration, 2014). For dichotomous outcomes, we used the odds ratios. We did not assess publication bias / small studies effect because fewer than 10 studies were included. Where data were missing, study authors were contacted.

## Results

We screened titles and abstracts of 898 articles including 14 registered trials and assessed 29 full text articles for inclusion (Figure 1). Main reasons for exclusion were lack of primary data and non-clinical setting. Full list of excluded studies with reasons can be found in the Supplementary file. Five published observational studies (in 6 reports) from 4 countries were included in the quantitative analysis^5,10–14^ (Table 1). Al Mohajer et al^10^ is a published full version of Hemmige et al^12^, with more complete data and therefore, was included in the analysis.

**Table 1:** Characteristics of included studies (n=5)

| Study ID, Area, Country | Study type and setting | Intervention period | Population | Intervention description | Outcomes reported |
| --- | --- | --- | --- | --- | --- |
| **Al Mohajer 2021<sup>10</sup>**<br>Texas, USA | Before-after; hospital | Jul 6 - Sep 7, 2020 | n=6527 HCPs, | Face shields (Lazarus 3D, Corvallis, OR, USA). Goggles allowed as an alternative for those unable to tolerate face shields. | n (%) HCPs tested positive; total patient days; hospital-acquired infections |
| **Bhaskar 2020<sup>5</sup>**<br>Chennai, India | Before-after; community screening project | May 10 - Jun 30, 2020 | n=62 HCPs | Face shields made of polyethylene terephthalate (250-µm thickness) | n (%) HCPs tested positive; n (%) community dwelling people visited + |
| **Hamilton 2021<sup>11</sup>**<br>Devon, UK | Before-after; District hospital COVID ward | mid-Dec 2020 - Feb 2021 | n=410 HCPs | Universal visors for all patient care | n (%) HCPs tested positive |
| **Rodriguez-Lopez 2021<sup>13</sup>**<br>Cali, Columbia | Case-control; hospital | Jun 10 - Jul 25, 2020 | n=223 HCWs | Face shield or goggles | Factors associated with SARS-CoV-2 infection including use of PPE, and compliance |
| **Shah 2021<sup>14</sup>**<br>Minnesota, USA | Retrospective cohort; Clinic | May 13 - Nov 30, 2020 | n=345 HCPs | Eye protection (face shield, protective wraparound eye wear, or a polycarbonate face shield or helmet) | n (%) HCPs tested positive |

HCPs – Healthcare professional, HCWs – Healthcare workers.

**Table 2:** Risk of bias in included studies assessed by ROBINS-I.

| Risk of bias domains<br>Study ID | Bias due to confounding | Bias in selection of participants | Bias in classification of interventions | Bias due to deviations from intended interventions | Bias due to missing data | Bias in measurement of outcomes | Bias in selection of the reported result | Overall risk of bias |
| --- | --- | --- | --- | --- | --- | --- | --- | --- |
| Al Mohajer 2021 | Moderate | Low | Low | Low | NI | Low | Low | Moderate |
| Bhaskar 2020 | NI | Low | Low | Low | Low | Low | Low | Moderate |
| Hamilton 2021 | Moderate | Low | Low | Low | NI | Low | Low | Moderate |
| Rodriguez-Lopez 2021 | Serious | Low | Low | NI | Low | Low | Low | Serious |
| Shah 2021 | Serious | Low | Low | NI | Low | Low | Low | Serious |

NI – No information

Description of the eye protections ranged from wraparound eyewear and goggles to full face shield or visor in addition to approved masks and other infection control measures in the corresponding clinical setting (Table 1). Three of the studies instituted the eye protection intervention at the time of peak infection in the respective communities^10,11,14^. However, none of them adjusted for change in risk of infection (e.g., community rates).

The high heterogeneity between studies precluded a meaningful meta-analysis. Forest plot of each study outcomes are presented in Figure 2. Most of the studies reported reduced number of infections after instigating eye protection for patient interaction. The three before-and-after studies all showed statistically significant and substantial reductions in SARS-CoV-2 infections favouring eye protection with odds ratios ranging from 0.04 to 0.6, corresponding to risk reductions of 96% to 40%^5,10,11^. However, the one case-control study reported odds ratio favouring no eye protection (OR 1.7, 95% CI 0.99, 3.0)^13^. None of the studies adjusted for potential confounders such as other protective behaviours.

**Figure 2.**
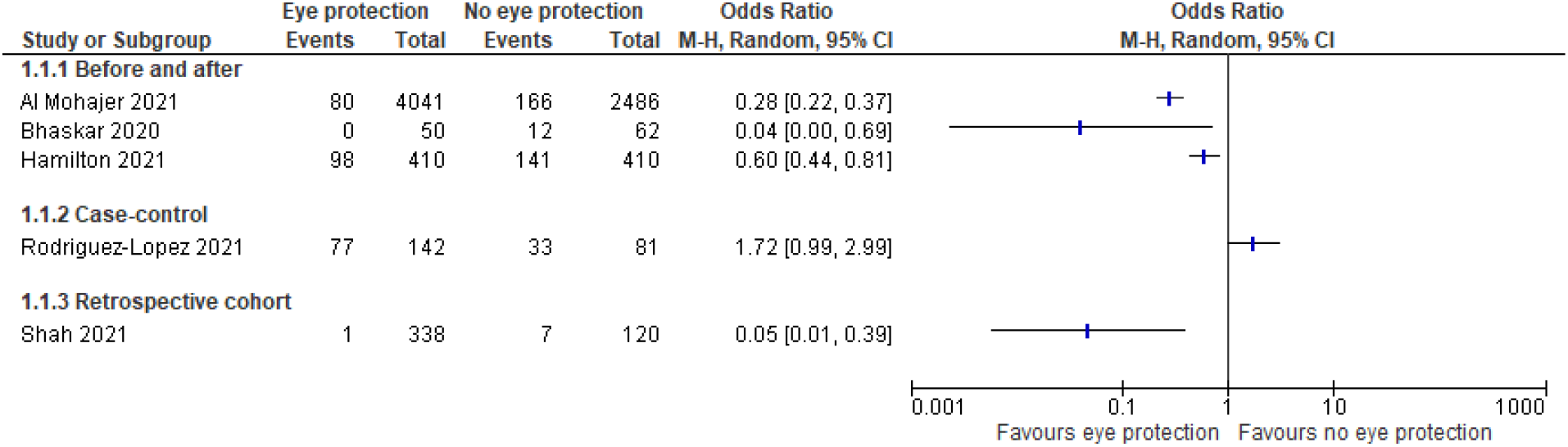
Forest plot of effect of eye protection in healthcare professionals.

**Figure 2:**
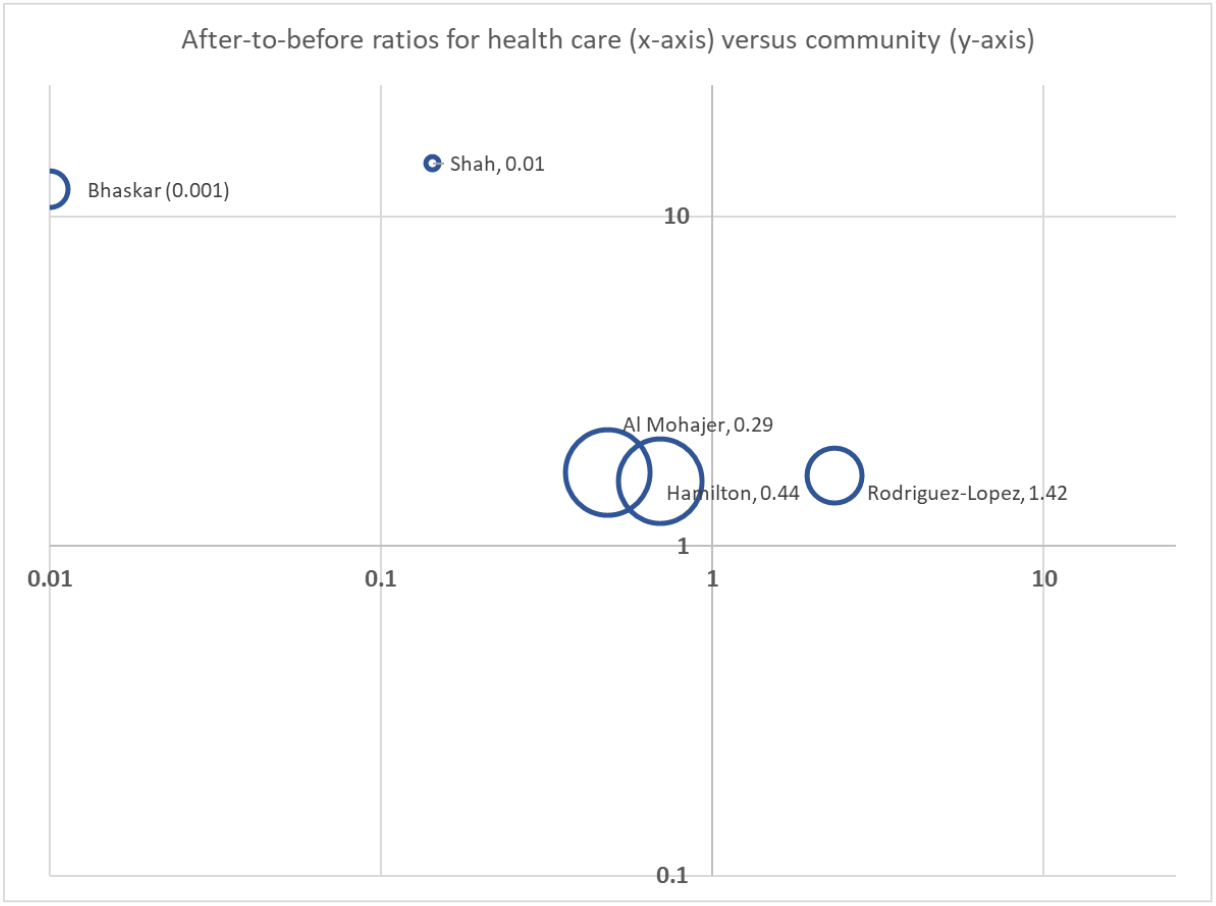
After-to-before ratios for health care (x-axis) versus community (y-axis) – studies are labelled together with the ratio of the two ratios.

We also collated data on community transmission in the same region as the studies from the local health authority reports and these all showed increases in cases. On the assumption that the denominators (numbers in health care facility and community) remained constant, we calculated the ratios of after-to-before for health care workers and compared these to after-to-before for the community (Figure 3). The reductions in health care worker infections after eye protection were not explained by changes in the community rates.

Figure 3 shows that for all studies the community rates of Covid19 increased in the “after” period compared to the before period (all are in the upper half of the figure above the x axis). Hence adjustments for the community rates would have further increased the apparent protective effects of eye protection. An adjustment of increase in community transmission in the Rodriguez-Lopez^13^ study would have reduced the case-control odds ratio of 1.72 to 1.42.

We also found three prevalence studies that looked at potential protective effect of regular eyewear in general population^15–17^. They suggested that wearing regular eyewear for more than 8 hours a day could reduce SARS-CoV-2 infection, but there was insufficient data to establish causation.

## Risk of bias

The risk of bias of the included studies was assessed by ROBINS-I for observational studies. The risk of bias within individual studies was judged overall as moderate to serious with confounding being the primary source (see Methods). Risk of selection bias, classification of intervention, measurement of outcomes and selection of reported results were judged to be low risk of bias.

## Discussion

Of the five observational studies identified, four showed substantial and statistically significant reductions in COVID-19 infections of health care workers after mandatory eye protection – mainly from face shields - was introduced; one case-control study showed an increase which was partly explained by an increase in community transmission. All five studies were non-randomized, and did not adjust for potential confounders, so the overall risk of bias was high. Therefore, the evidence summarised here is very low certainty.

One important confounder for the before-after studies is any change in community transmission between the before-after periods; as demonstrated in Figure 2, the community rates were generally higher in the after period; hence adjustment would only increase the size of the estimated reduction. However, the higher rates may also mean increases in other protective behaviours which are not reported in any of the studies. Finally, while the studies’ main intervention was face shields, they also allowed the use of some other forms of eye protection such as goggles.

A previous review of observational studies on the effects in of eye protection in the SARS and MERS epidemics found a reduction in transmission of 66% and 76% respectively^3^. These reductions are comparable with the reductions seen in the three before-after studies of this review. Several laboratory studies using mannikins have examined the potential effects of face shields but vary greatly in their design and their application to real-world settings. One study tested facing mannikins 25cm apart with the emitter sending an aerosol spray with particles with a range of size from less than 0.3µm to 10 µm; they found a reduction in particles received of 55% with face shields compared to 22% for a mask, and 97% for both^18^. A study with a mannikin 60cm from the spray found greater reductions and face shields providing greater protection than masks^19^. However, these studies were of water aerosols not transmission of viral particles and are very incomplete simulations of human interaction. Neither separated eye protection specifically from face protection.

These studies provide suggestive evidence that face shields provide some protective effect, and that this may be substantial. These studies cannot determine how much of the protective effect is due to reduction of transmission from the eyes via nasolacrimal duct to nose. Furthermore, a face shield – the main protection used – may provide additional inhalation protection as seen in some of the laboratory studies. While goggles also provide eye protection, face shields will likely give substantial protection against inhalation of droplets as well as eye protection and are more comfortable to wear. Hence face shields - in addition to masks - should be considered for higher risk situations – such as contact tracing, quarantine workers, and some primary care consultations - or when there is substantial Covid spread in the community. Additional protection is likely to be particularly important to health care workers in settings where currently only face masks are being used.

While these observational studies show an interesting potential protection from face shields as add-on to face masks, they do not clarify whether such protection is from reduced inhalation or eye protection. Trials of the incremental value of face shields in addition or instead of face masks and comparative studies of face shields and eye goggles with face masks all seem warranted. Such studies should also measure comfort and adherence of different options used, as correct and sustained usage is also critical to effectiveness.

## Data Availability

All supporting data are available and provided in the Supplementary material.

## Supplementary material

**Supplement 1. Full search strategy PubMed:**

(“COVID-19” [Mesh] OR “SARS-CoV-2” [Mesh] OR “COVID-19” [Supplementary Concept] OR “SARS-CoV-2 variants” [Supplementary Concept] OR “COVID-19” [tiab] OR COVID19[tiab] OR “COVID 19” [tiab] OR “SARS-CoV-2” [tiab] OR “2019-nCoV” [tiab] OR “Novel coronavirus” [tiab] OR “Coronavirus 2019” [tiab] OR “Coronavirus 19” [tiab] OR “COVID 2019” [tiab] OR “2019 ncov” [tiab] OR “Wuhan coronavirus” [tiab])

AND

(“Eye Protective Devices” [Mesh] OR Glasses[tiab] OR Goggle[tiab] OR “Eye protection” [tiab] OR Faceshield[tiab] OR Faceshields[tiab] OR Goggles[tiab] OR “Face shield” [tiab] OR “Face shields” [tiab] OR Visors[tiab] OR “Prophylactic measures” [tiab])

AND

(“transmission” [sh] OR Prevention[tiab] OR Reduces[tiab] OR Spread[tiab] OR Efficacy[tiab] OR Reduction[tiab] OR Transmission[tiab])

AND

(“randomized controlled trial” [pt] OR “controlled clinical trial” [pt] OR randomized[tiab] OR randomised[tiab] OR placebo[tiab] OR randomly[tiab] OR trial[tiab] OR groups[tiab] OR Crossover[tiab] OR “Comparative Study” [pt] OR “Evaluation Study” [pt] OR “Epidemiologic Studies” [Mesh] OR “case-control studies” [Mesh] OR “Cohort Studies” [Mesh] OR “case control” [tiab] OR Cohort[tiab] OR “Follow up” [tiab] OR Observational[tiab] OR Longitudinal[tiab] OR Prospective[tiab] OR retrospective[tiab] OR “cross sectional” [tiab] OR “Cross-Sectional Studies” [Mesh] OR Investigated[tiab] OR Evaluated[tiab] OR Impact[tiab] OR Analysis[tiab] OR Statistics[tiab] OR Data[tiab] OR “statistics and numerical data” [sh] OR “epidemiology” [sh] OR Experimental[tiab] OR Experiment[tiab] OR “Letter” [pt] OR “Comment” [pt])

### Embase

(‘coronavirus disease 2019’/exp OR ‘Severe acute respiratory syndrome coronavirus 2’/exp OR COVID-19:ti,ab OR COVID19:ti,ab OR “COVID 19”:ti,ab OR SARS-CoV-2:ti,ab OR 2019-nCoV:ti,ab OR “Novel coronavirus”:ti,ab OR “Coronavirus 2019”:ti,ab OR “Coronavirus 19”:ti,ab OR “COVID 2019”:ti,ab OR “2019 ncov”:ti,ab OR “Wuhan coronavirus”:ti,ab)

AND

(“eye protective device” /exp OR Glasses:ti,ab OR Goggle:ti,ab OR “Eye protection”:ti,ab OR Faceshield:ti,ab OR Faceshields:ti,ab OR Goggles:ti,ab OR “Face shield”:ti,ab OR “Face shields”:ti,ab OR Visors:ti,ab OR “Prophylactic measures”:ti,ab)

AND

(Prevention:ti,ab OR Reduces:ti,ab OR Spread:ti,ab OR Efficacy:ti,ab OR Reduction:ti,ab OR Transmission:ti,ab)

AND

(random* OR factorial OR crossover OR placebo OR blind OR blinded OR assign OR assigned OR allocate OR allocated OR ‘crossover procedure’/exp OR ‘double-blind procedure’/exp OR ‘randomized controlled trial’/exp OR ‘single-blind procedure’/exp OR ‘epidemiology’/exp OR ‘controlled study’/exp OR ‘cohort analysis’/exp OR “case control”:ti,ab OR Cohort:ti,ab OR “Follow up”:ti,ab OR Observational:ti,ab OR longitudinal:ti,ab OR Prospective:ti,ab OR retrospective:ti,ab OR “cross sectional”:ti,ab OR ‘Cross-Sectional Studies’/exp OR Investigated:ti,ab OR Analysis:ti,ab OR Statistics:ti,ab OR Data:ti,ab)

### Cochrane CENTRAL

([mh “COVID 19”] OR [mh “SARS CoV 2”] OR “COVID 19”:ti,ab OR COVID19:ti,ab OR “COVID 19”:ti,ab OR “SARS CoV 2”:ti,ab OR 2019 nCoV:ti,ab OR “Novel coronavirus”:ti,ab OR “Coronavirus 2019”:ti,ab OR “Coronavirus 19”:ti,ab OR “COVID 2019”:ti,ab OR “2019 ncov”:ti,ab OR “Wuhan coronavirus”:ti,ab)

AND

([mh “Eye Protective Devices”] OR Glasses:ti,ab OR Goggle:ti,ab OR “Eye protection”:ti,ab OR Faceshield:ti,ab OR Faceshields:ti,ab OR Goggles:ti,ab OR “Face shield”:ti,ab OR “Face shields”:ti,ab OR Visors:ti,ab OR “Prophylactic measures”:ti,ab)

AND

([mh /TM] OR Prevention:ti,ab OR Reduces:ti,ab OR Spread:ti,ab OR Efficacy:ti,ab OR Reduction:ti,ab OR Transmission:ti,ab)

### Cochrane COVID-19 Study Register

(“Eye protection” OR Faceshield OR Faceshields OR “Face shield” OR “Face shields”) AND

(Prevention OR Reduces OR Spread OR Efficacy OR Reduction OR Transmission)

### Europe PMC (Preprints)

(COVID-19 OR SARS-CoV-2 OR COVID-19 OR COVID19 OR “COVID 19” OR SARS-CoV-2 OR 2019-nCoV OR “Novel coronavirus” OR “Coronavirus 2019” OR “Coronavirus 19” OR “COVID 2019” OR “2019 ncov” OR “Wuhan coronavirus”)

AND

(“Eye protection” OR Faceshield OR Faceshields OR “Face shield” OR “Face shields”) AND

(Prevention OR Reduces OR Spread OR Efficacy OR Reduction OR Transmission)

AND

(SRC:PPR)

**Supplement 2.** Excluded studies following full text screening with reasons.

| No. | Excluded articles | Reason |
| --- | --- | --- |
| 1 | Chua JX, Ong L, Tan CH. Innovative Face Shields Help Frontliners Face-off COVID-19 Pandemic. *Ann Acad Med Singap.* 2020;49(12):1044-7. | No primary data |
| 2 | Gupta S, Jangra RS, Gupta S, Gujrathi AV, Sharma A. Makeshift face shield for healthcare professionals during the COVID-19 pandemic. *Clin Exp Dermatol.* 2020;45(6):751-2. | No primary data |
| 3 | Ha JF. The COVID-19 pandemic and face shields. *Br J Surg.* 2020;107(10):e398. | No primary data |
| 4 | Khunti K, Adisesh A, Burton C, Chan XHS, Coles B, Durand-Moreau Q, et al. Analysis ;The efficacy of PPE for COVID-19-type respiratory illnesses in primary and community care staff. *British Journal of General Practice.* 2020;70(697):413-6. | No primary data |
| 5 | MacIntyre CR, Wang Q. Physical distancing, face masks, and eye protection for prevention of COVID-19. *Lancet* 2020; 395(10242): 1950-1. | No primary data |
| 6 | Marra AR, Edmond MB, Popescu SV, Perencevich EN. Examining the need for eye protection for coronavirus disease 2019 (COVID-19) prevention in the community. *Infect Control Hosp Epidemiol* 2021; 42(5): 646-7. | No primary data |
| 7 | Hawkins ES, Fertel BS, Muir MR, Meldon SW, Delgado FJ, Smalley CM. Adding eye protection to universal masking reduces COVID-19 among frontline emergency clinicians to the level of community spread. *Am J Emerg Med* 2020. | No primary data/insufficient detail |
| 8 | Tan Gse LKZSMMLVSCMPBFNOTABSLY. Effect of extended use N95 respirators and eye protection on personal protective equipment (PPE) utilization during SARS-CoV-2 outbreak in Singapore. *Antimicrobial resistance and infection control* 2020; 9(1): 86. | No primary data |
| 9 | Ha JF, Collier R, Kop AM, Morrison DA. The COVID-19 pandemic and ENT modified face shields. *Br J Surg* 2020; 107(11): e448-e9. | Lab study |
| 10 | Li L, Niu M, Zhu Y. Assessing the effectiveness of using various face coverings to mitigate the transport of airborne particles produced by coughing indoors. *Aerosol Science and Technology* 2020; 55(3): 332-9. | Lab study |
| 11 | Lindsley WG, Blachere FM, Law BF, Beezhold DH, Noti JD. Efficacy of face masks, neck gaiters and face shields for reducing the expulsion of simulated cough-generated aerosols. *Aerosol Science and Technology* 2021; 55(4): 449-57. | Lab study |
| 12 | Ronen A, Rotter H, Elisha S, et al. Investigation of the protection efficacy of face shields against aerosol cough droplets. *J Occup Environ Hyg* 2021; 18(2): 72-83. | Lab study |
| 13 | Schmitt J, Jones L, Aeby E, Gloor C, Moser B, Wang J. Protection Level and Reusability of a Modified Full-Face Snorkel Mask as Alternative Personal Protective Equipment for Healthcare Workers During the COVID-19 Pandemic. 2020. | Lab study |
| 14 | Stephenson T, Cumberland C, Kibble G, et al. Evaluation of Facial Protection Against Close-Contact Droplet Transmission. 2021. | Lab study |
| 15 | Tretiakow D, Tesch K, Skorek A. Mitigation effect of face shield to reduce SARS-CoV-2 airborne transmission risk: Preliminary simulations based on computed tomography. Environmental Research 2021; 198. | Lab study |
| 16 | Verma S, Dhanak M, Frankenfield J. Visualizing droplet dispersal for face shields and masks with exhalation valves. 2020. | Lab study |
| 17 | Viola IM, Peterson B, Pisetta G, et al. Face Coverings, Aerosol Dispersion and Mitigation of Virus Transmission Risk. 2021. | Lab study |
| 18 | Wendling JM, Fabacher T, Pébaÿ PP, Cosperec I, Rochoy M. Experimental Efficacy of the Face Shield and the Mask against Emitted and Potentially Received Particles. Int J Environ Res Public Health 2021; 18(4). | Lab study |
| 19 | Lehrer S, Rheinstein P. Eyeglasses Reduce Risk of COVID-19 Infection. In Vivo 2021; 35(3): 1581-2. | Regular eyewear study |
| 20 | Zeng W, Wang X, Li J, et al. Association of Daily Wear of Eyeglasses With Susceptibility to Coronavirus Disease 2019 Infection. JAMA Ophthalmol 2020; 138(11): 1196-9. | Regular eyewear study |
| 21 | Saxena AK. Risk of Corona virus disease 2019 (COVID-19) among spectacles wearing population of Northern India. 2021. | Regular eyewear study |
| 22 | Nct. Effectiveness and Adherence to Closed Face Shields to Prevent COVID-19 Transmission.<br>https://clinicaltrials.gov/show/NCT04647305, 2020.<br>https://www.cochranelibrary.com/central/doi/10.1002/central/CN-02205986/full | On-going trial with no results |
| 23 | Nct. Enhanced Hood PPE to Minimize COVID-19 Transmission to Front-line Health Care Workers.<br>https://clinicaltrials.gov/show/NCT04373096, 2020.<br>https://www.cochranelibrary.com/central/doi/10.1002/central/CN-02103676/full | On-going trial with no results |

